# Evaluation of sleep pattern due to stress in undergraduate medical students and its impact on health and academic performance: A cross-sectional study from tertiary health center of Central India

**DOI:** 10.1101/2021.09.19.21263802

**Authors:** Kusum Gandhi, Yashvardhan Godaria, G Revadi

**Affiliations:** Department of Anatomy, AIIMS Bhopal; M.B.B.S student, AIIIMS Bhopal; Department of Community & Family Medicine AIIMS Bhopal

**Keywords:** sleep pattern, medical students, academic stress factors, coping strategies

## Abstract

**Background:** Good quality sleep is essential for good health and well-being. Medical students are at no exception to this and are prone to greater risk for sleep deprivation. The major reason being challenges to maintain a high level of academic achievement and constant thirst to acquire new learning skills and knowledge. However, in this process they are circumstanced to various levels of stress that might cause potential damage to their cognitive functioning and mental exhaustion to a certain extent.

**Objectives:** Thus, our study objectives were to evaluate the sleep pattern in first- and second-year medical students. To understand how the stress levels and academic performance are related to sleep pattern and to explore the copying strategies of stress in our study participants.

**Methodology:** This cross-sectional study was conducted using a self-reported, web-based, questionnaire that included questions on sleep quality and deprivation through Pittsburgh sleep quality index. All the eligible students of first and second year who were part of a premiere teaching hospital during February and March 2021 were included. Data was analysed using IBM SPSS version 24.

**Results:** Out of 180 participants, 91(50.55%) had their initiation of sleeping time from 12-2 am and also, majority of students 112 (62.22%) had a sleep duration of six to eight hours. However, 119 (66.1%) students had self-reported change in sleeping pattern which was found to be significantly associated with relatively greater number of academic factors as compared to social factors. Most of the students scored between 50-60% score in their four assessments amongst which their first assessment was significantly associated with change in sleep pattern (P 0.040). Also, these individual assessment score was found to significantly affect their duration of sleep. The common coping strategies adopted by students under study were talking to family members/ friends, music/ book reading (hobby).

**Conclusion:** Majority of students in our study had reported change in sleeping pattern. Also, association between stress factors and change in sleeping pattern were observed with academic stress factors proving to be more significantly associated than social stress factors. The academic performance of students was also found to be associated with change in sleeping pattern and duration of sleep.

## Introduction

Thomas Dekkar has well documented that “Sleep is the golden chain that ties health and our bodies together”.^1^ Sleep is defined as a recurring, reversible neuro-behavioral state of relative perceptual disengagement from and unresponsiveness to the environment.^2^ Sleep is typically accompanied (in humans) by postural recumbence, behavioral quiescence, and closed eyes.^2^ Wolfson and Carskadon (2003) described that main correlates of poor academic performance are self-reported erratic sleep wake schedule, short total sleep time, phase delay, and poor-quality sleep.^3^ Good quality sleep is essential for good health and well-being. However, lifestyle and environmental factors are increasingly causing difficulties in sleeping. Sleep disturbance is frequently considered the most serious consequence of environmental noise by WHO.^4^

The main effects of sleep deprivation include physical effects (sleepiness, fatigue, hypertension) cognitive impairment and mental health complications. Such inadequate rest impairs the ability to think, to handle stress, to maintain a healthy immune system, and to moderate emotions (WHO).^4^ Considerably more sleep is required for the teenagers, to perform, optimally than younger children or adults. Low academic performance has been found to be significantly more common in students with insomnia and daytime sleepiness.^5^ Four fundamental characteristics of sleep like sleep quantity, sleep quality, sleep regularity, and sleep phase scheduling influence academic performance.^6^

It is believed that the medical students are at greater risk for sleep deprivation.^7^ The main causes being challenges to maintain a high level of academic achievement, trying to acquire adequate medical knowledge, learning clinical skills and adjusting to an ever-changing hospital environment in a limited period. High level of stress may have a negative effect on cognitive functioning and learning of students in the medical school.^8^ The majority of studies on stress in medical education focuses on the documentation of stress and information on the correlates of stress.^9,10^ It is not just the undergraduate study period which brings stress but it may continue during the internship, postgraduate study period, and later into physician’s practical life.^11,12,13^ The stress may also reach burn out levels.^14^ Thus, our study was conducted with the objectives to evaluate the pattern of sleep-in first- and second-year medical students. To understand how the stress levels and academic performance are related to sleep pattern and to explore the copying strategies of stress in our study participants.

### Methodology

This cross-sectional study was conducted among all the eligible 200 first- and second-year medical students, from a premier tertiary care teaching hospital of central India for a 2 months period (February and March 2021). Sample size was calculated using Sample size calculators with reference study: “Study of sleep patterns and sleep problems of undergraduate students from different professional courses” by Modi et.al. With 95% Confidence interval, alpha error as and standard deviation of 17, the estimated sample size was 178.

A web-based self-administered questionnaire was developed using the Google forms, which is free for non-commercial use. The structured questionnaire was developed in English language containing closed ended questions related to sleep quality and sleep deprivation according to PSQI (Pittsburgh sleep quality index). The questionnaire was derived from the medical students stressor questionnaire for analysis of the stress levels in the students. A link to the questionnaire, along with the electronic copies of participant information sheet and a copy of consent form were shared individually with each one of them on WhatsApp (Facebook Corp), which is a mobile messaging application. At least two reminders were sent to students non-responders through phone call, WhatsApp or personal contact before marking him/her as non-responder. Clearance from the institutional human ethics committee of AIIMS Bhopal was obtained (IHEC: 2020/STS/Jan/01) prior to start of study.

The following operational definitions were used in the present study:

1. Usual time of sleeping (bedtime) in the past 28 days.
2. Change of self-perceived sleeping pattern in the last 28 days.
3. Use of any medications for sleep initiation in the last 28 days.

### Statistical analysis

Data was entered in Microsoft Excel 2010 and analysis was performed using Statistical Package for Social Sciences version 24 (IBM SPSS). Proportions and median with interquartile range were calculated, based on the non-parametric distribution appropriately. Chi-square test was used to compare proportions among groups. Mann Whitney U test was applied to assess the significance between dependent categorical variable and independent continuous variables. *P* value <0 05 was significant.

## Results

There were 180 first- and second-year medical undergraduate students who participated in the study. Firstly, we tried to understand the sleep pattern of our study participants in terms of initiation of sleeping time, sleep duration and self-perceived change in sleeping pattern and use of medications if any. In our study most students i.e., 91 (50.55%) had their initiation of sleeping time from 12-2 am, 69 (38.3%) slept before 12 am while 20 (11.10%) slept after two am. Also, majority of students 112 (62.22%) had a sleep duration of six to eight hours as compared to 48 (28.66%) and 20 (11.10%) students who had their duration of sleep less than six hours and more than eight hours respectively. However, 119 (66.1%) students self-reported for change in sleeping pattern in recent times. It was also found that only 6 (3.33%) were using medications for sleep.

Secondly, we enlisted 25 core factors of academic stress and 16 factors for that of social stress as given in Supplementary File 1. Students were asked to number the responses as 0,1,2,3,4 where 0 corresponds to - no stress, 1 - mild stress, 2 - moderate stress, 3 - high stress, 4 - severe stress. Amongst the 25 academic factors, the most common factors accounting for severe stress as given in Supplementary Table 1 were large amount of content to be learnt, lack of time to review what has been learnt and heavy workload. Amongst the social factors shown in Supplementary Table 2, those factors accounting for severe stress were poor motivation to learn, lack of time for friends and lack of time for family.

The scores for individual academic stress factors as median (IQR) were tabulated against the dependent variable, change in sleep pattern (Yes/No) in Table 1. Following Mann Whitney test, it was found that the following academic stress factors were found to be significantly associated with their self -reported change in sleep patterns. Those factors includes: failing behind in dissection in anatomy (P 0.011), large amount of content to be learnt (P 0.006), lack of time to review what has been learnt (P 0.01), unable to answer the questions from the teacher (P 0.009), feeling of incompetence (P 0.007), not enough feedback / guidance / encouragement from teacher (P 0.009), not enough study material (P 0.008), language barrier (P 0.007) and education system (P 0.003)

**Table 1:** Relationship between academic stress factors and change in sleeping pattern:

| Academic stress factors | Change in sleeping pattern<br>Median (IQR) |  | P value |
| --- | --- | --- | --- |
|  | Present | Absent |  |
| Assessment / examinations | 2 (1-3) | 2 (1-2) | 0.053 |
| Failing behind in dissection in anatomy | 2 (0-3) | 1 (0-2) | 0.011 |
| Large amount of content to be learnt | 3 (2-3) | 2 (1-3) | 0.006 |
| Lack of time to review what has been learnt | 2 (2-3) | 2 (1-3) | 0.01 |
| Heavy workload | 2 (1-3) | 2 (1-3) | 0.040 |
| Difficulty understanding the content | 2 (1-3) | 1 (0-2) | 0.040 |
| Learning context full of competition | 1 (1-3) | 1 (0-2) | 0.109 |
| Unable to answer the questions from the teacher | 2 (1-3) | 1 (1-2) | 0.009 |
| Unjustified grading process | 1 (0-3) | 1 (0-2) | 0.080 |
| Getting poor marks | 2 (1-3) | 1 (1-2) | 0.228 |
| Participation in class presentation/discussion | 1 (0-2) | 1 (0-2) | 0.066 |
| Need to do well (imposed by others) | 1 (0-3) | 1 (0-2) | 0.045 |
| Feeling of incompetence | 1 (0-3) | 1 (0-2) | 0.007 |
| Unwillingness to study medicine | 0 (0-2) | 0 (0-1) | 0.331 |
| Parental wish for you to study medicine | 0 (0-1) | 0 (0-1) | 0.171 |
| Not enough feedback /guidance<br>/encouragement from teacher | 1 (0-2) | 0 (0-1) | 0.009 |
| Uncertainty of what is expected of me | 1 (0-2) | 1 (0-2) | 0.060 |
| Lack of recognition for work done | 1 (0-2) | 1 (0-1) | 0.177 |
| Inappropriate assignments | 1 (0-2) | 1 (0-2) | 0.213 |
| Lack of teaching skill in teacher | 1 (0-2) | 0 (0-1) | 0.082 |
| Not enough study material | 0 (0-2) | 0 (0-1) | 0.008 |
| Lack of relevance of the course in real life | 1 (0-2) | 0 (0-1) | 0.092 |
| Language barrier | 0 (0-1) | 0 (0-0) | 0.007 |
| Lack of self-assessment | 1 (0-2) | 1 (0-2) | 0.106 |
| Education system | 1 (0-3) | 1 (0-2) | 0.003 |

Similarly in table 2, the scores of social factors were assessed for association with the self-reported change in sleep pattern. There were only two social stress factors, financial issues (P 0.025) and health issues (P 0.036) which were significantly associated with change in sleep pattern following Mann Whitney test.

**Table 2.**
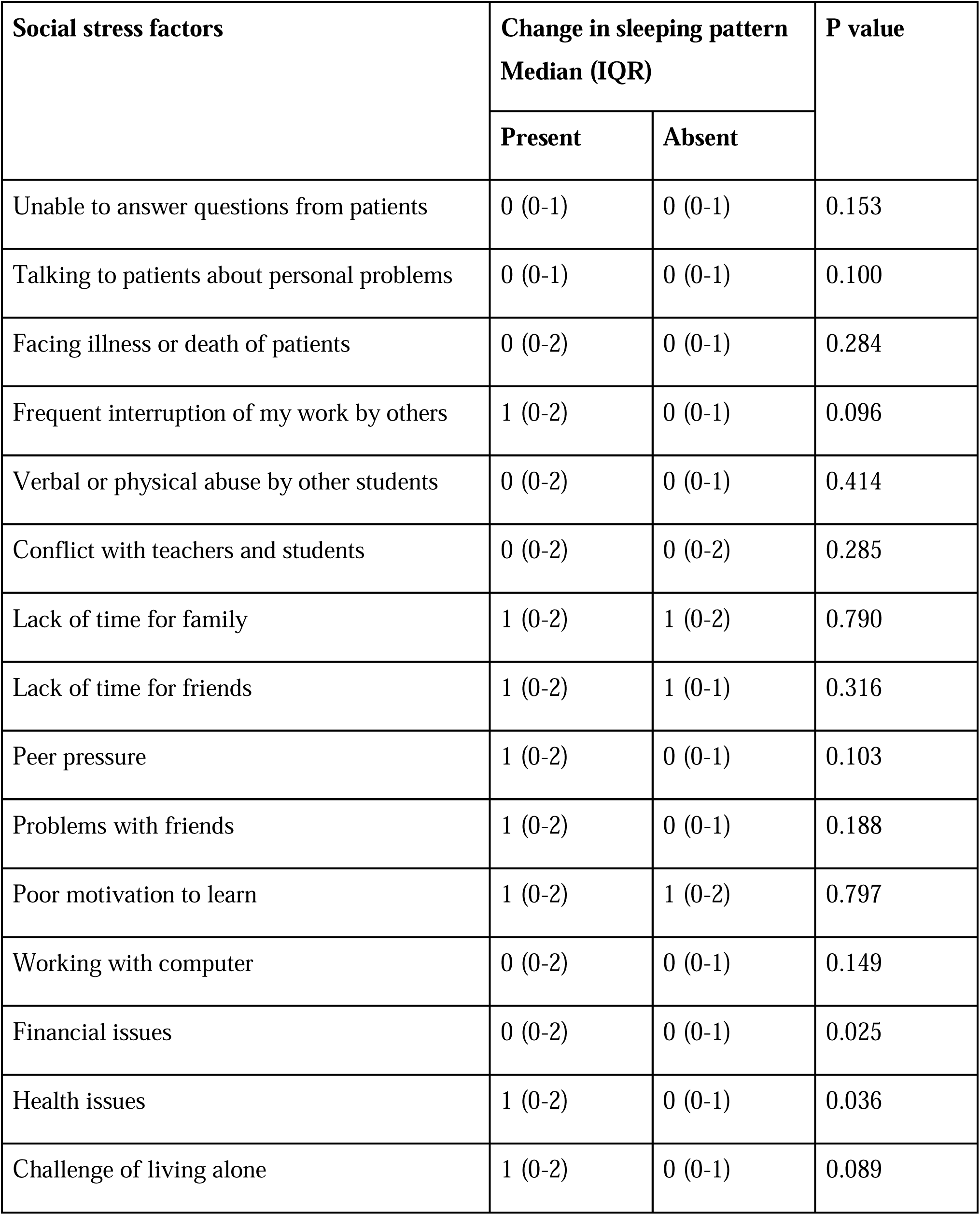
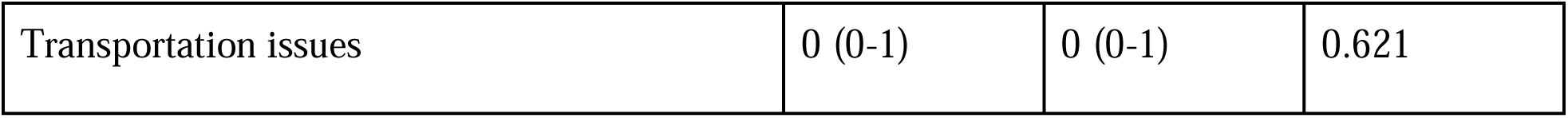
Relationship between social stress factors and change in sleeping pattern.

| **Social stress factors** | **Change in sleeping pattern**<br>**Median (IQR)** |  | **P value** |
| --- | --- | --- | --- |
|  | **Present** | **Absent** |  |
| Unable to answer questions from patients | 0 (0-1) | 0 (0-1) | 0.153 |
| Talking to patients about personal problems | 0 (0-1) | 0 (0-1) | 0.100 |
| Facing illness or death of patients | 0 (0-2) | 0 (0-1) | 0.284 |
| Frequent interruption of my work by others | 1 (0-2) | 0 (0-1) | 0.096 |
| Verbal or physical abuse by other students | 0 (0-2) | 0 (0-1) | 0.414 |
| Conflict with teachers and students | 0 (0-2) | 0 (0-2) | 0.285 |
| Lack of time for family | 1 (0-2) | 1 (0-2) | 0.790 |
| Lack of time for friends | 1 (0-2) | 1 (0-1) | 0.316 |
| Peer pressure | 1 (0-2) | 0 (0-1) | 0.103 |
| Problems with friends | 1 (0-2) | 0 (0-1) | 0.188 |
| Poor motivation to learn | 1 (0-2) | 1 (0-2) | 0.797 |
| Working with computer | 0 (0-2) | 0 (0-1) | 0.149 |
| Financial issues | 0 (0-2) | 0 (0-1) | 0.025 |
| Health issues | 1 (0-2) | 0 (0-1) | 0.036 |
| Challenge of living alone | 1 (0-2) | 0 (0-1) | 0.089 |
| Transportation issues | 0 (0-1) | 0 (0-1) | 0.621 |

Thirdly, students were asked to fill the approximate marks obtained in the previous four academic assessments. It was evident from Supplementary Table 3 that most students scored between 50-60% in all the assessments while few students scored in 71-80% in all the assessments. The association between individual assessment scores and self-reported change in sleep pattern were tabulated in Table 3. It was found that only the first assessment performance (P 0.040) had significant relationship with the change in sleeping pattern following Chi square test.

**Table 3:** Relationship between Academic performance in sequential assessments and change in sleeping pattern.

| Score in assessment 1 | Change in sleeping pattern (Number of students) (Percentage of students in parentheses) |  | Total number of students | P value |
| --- | --- | --- | --- | --- |
|  | Present | Absent |  |  |
| <50 | 36 (77.5) | 11 (22.5) | 47 | 0.040 |
| 50-60 | 43 (71.7) | 17 (28.3) | 60 |  |
| 61-70 | 23 (63.9) | 13 (36.1) | 36 |  |
| 71-80 | 6 (37.5) | 10 (62.5) | 16 |  |
| >80 | 13 (56.5) | 10 (43.5) | 23 |  |
| Score in assessment 2 |  |  |  |  |
| <50 | 31 (77.5) | 9 (22.5) | 40 | 0.107 |
| 50-60 | 43 (71.7) | 17 (28.3) | 60 |  |
| 61-70 | 23 (56.1) | 18 (43.9) | 41 |  |
| 71-80 | 13 (68.4) | 6 (31.6) | 19 |  |
| >80 | 11 (50) | 11 (50) | 22 |  |
| Score in assessment 3 |  |  |  |  |
| <50 | 23 (74.2) | 8 (25.8) | 31 | 0.361 |
| 50-60 | 48 (72.7) | 18 (27.3) | 66 |  |
| 61-70 | 29 (60.4) | 19 (39.6) | 48 |  |
| 71-80 | 8 (57.1) | 6 (42.9) | 14 |  |
| >80 | 13 (66.5) | 10 (43.5) | 23 |  |
| Score in assessment 4 |  |  |  |  |
| <50 | 21 (70) | 9 (30) | 30 | 0.093 |
| 50-60 | 49 (75.4) | 16 (24.6) | 65 |  |
| 61-70 | 29 (67.4) | 14 (32.6) | 43 |  |
| 71-80 | 9 (52.9) | 8 (47.1) | 17 |  |
| >80 | 13 (48.1) | 14 (51.9) | 27 |  |

Similarly in table 4, duration of sleep was cross tabulated with the individual assessment scores using Chi-square assessment. Table 4 shows that the significant relationship between academic score and duration of sleep pattern using Chi square test in assessments 1 (P 0.003), 2 (P 0.048) and 4 (P 0.019) and borderline significant in assessment 3 (P 0.084). Furthermore, in all these four assessments, maximum percentage of students scored 50-60 (less than average score) and for these students, most common sleeping duration was 6-8 hours.

**Table 4.** Relationship between academic performance and quantity of sleep.

| Score in Assessment 1 | Duration of sleep |  |  | Total number of students | P value |
| --- | --- | --- | --- | --- | --- |
|  | <6 hours | 6-8 hours | >8 hours |  |  |
| <50 | 21 (44.7) | 25 (53.2) | 1 (2.1) | 47 | 0.003 |
| 50-60 | 16 (26.7) | 42 (70) | 2 (3.3) | 60 |  |
| 61-70 | 8 (22.2) | 22 (61.1) | 6 (16.7) | 36 |  |
| 71-80 | 1 (6.3) | 15 (93.8) | 0 (0) | 16 |  |
| >80 | 4 (17.4) | 18 (78.3) | 1 (4.3) | 23 |  |
| Score in assessment 2 |  |  |  |  |  |
| <50 | 18 (45.0) | 21 (52.5) | 1 (2.5) | 40 | 0.048 |
| 50-60 | 18 (30.0) | 40 (66.7) | 2 (3.3) | 60 |  |
| 61-70 | 7 (17.1) | 31 (75.6) | 3 (7.3) | 41 |  |
| 71-80 | 4 (21.1) | 12 (63.2) | 3 (15.8) | 19 |  |
| >80 | 3 (13.6) | 18 (81.8) | 1 (4.5) | 22 |  |
| Score in assessment 3 |  |  |  |  |  |
| <50 | 15 (48.4) | 15 (48.4) | 1 (3.2) | 31 | 0.084 |
| 50-60 | 20 (30.3) | 43 (65.2) | 3 (4.5) | 66 |  |
| 61-70 | 10 (20.8) | 35 (72.9) | 3 (6.3) | 48 |  |
| 71-80 | 1 (7.1) | 11 (78.6) | 2 (14.3) | 14 |  |
| >80 | 4 (17.4) | 18 (78.3) | 1 (4.3) | 23 |  |
| Score in assessment 4 |  |  |  |  |  |
| <50 | 12 (40.0) | 17 (56.7) | 1 (3.3) | 30 | 0.019 |
| 50-60 | 23 (35.4) | 40 (61.5) | 2 (3.1) | 65 |  |
| 61-70 | 9 (20.9) | 28 (65.1) | 6 (14.0) | 43 |  |
| 71-80 | 1 (5.9) | 16 (94.1) | 0 (0.0) | 17 |  |
| >80 | 5 (18.5) | 21 (77.8) | 1 (3.7) | 27 |  |

The supportive figure 1 displayed below shows, various copying strategies listed along the vertical axis and the responses of students for each of those coded as five different colours in horizontal axis below. It is more subjectively evident that most of these students had been “doing a little bit” (marked in green) of each of the listed copying strategies. The common coping strategies adopted by students were good sleep, talking to family members/ friends, music/ book reading (hobby).

**Figure 1:**
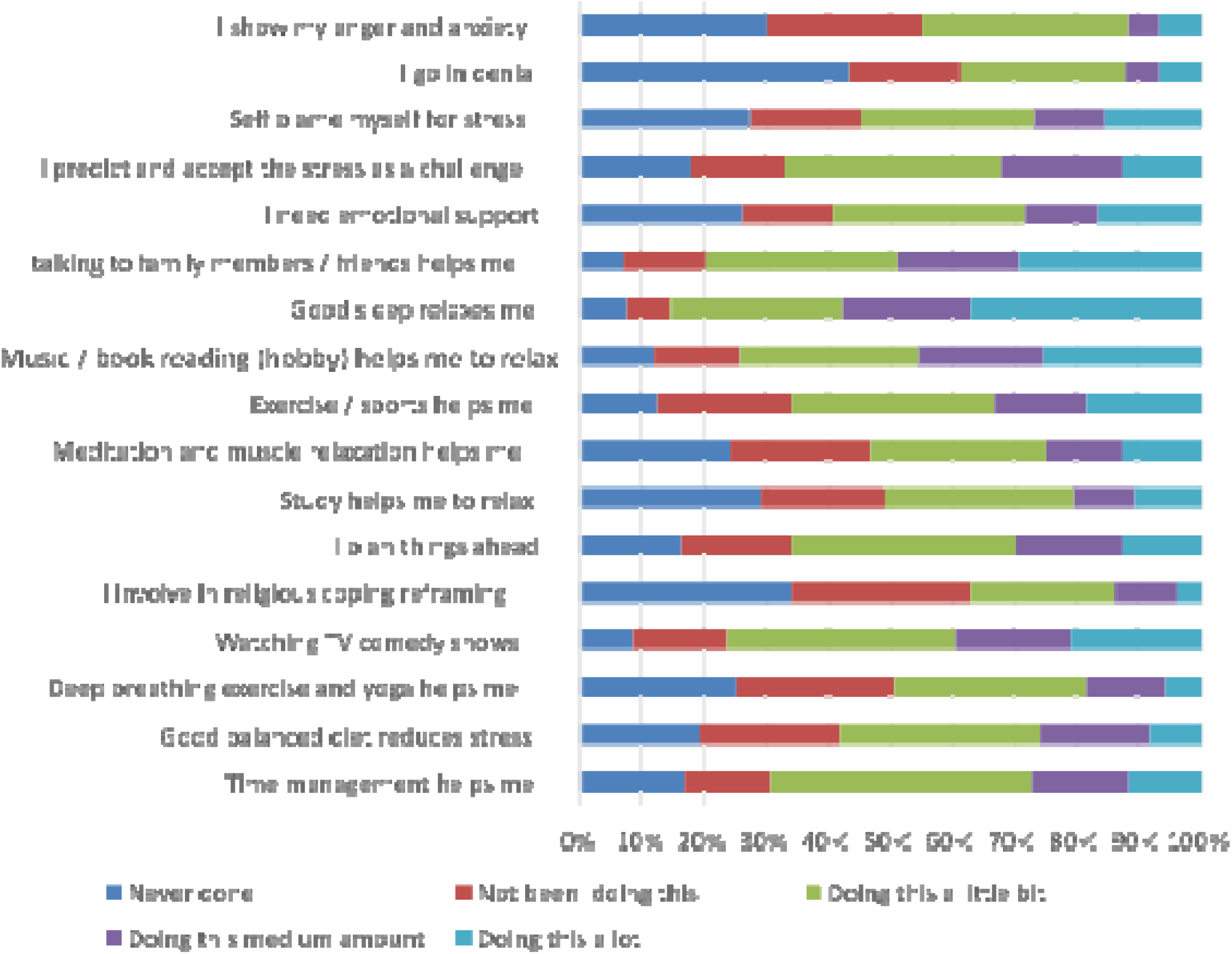
Copying strategies to control stress

## Discussion

More than half of the participants in this study had a sleep duration for six to eight hours. Hamed et al. (2015) in his study showed that nearly half of undergraduate medical students sleep in the same duration of six to eight hours daily.^15^ Also, two thirds of the students in our study had a change in sleeping pattern. Similar results like poor sleep-wake habits (in terms of hours slept, sleep quality, daytime sleepiness) were also reported among medical students in a study by Alsagaff et al. (2016).^16^ This finding to a certain extent in our study could be explained due to the influence of new college lifestyle and hostel environment.

Secondly, while trying to study the role of stress factors: it was found that the students were more stressed by academic factors than social factors that led to a significant change in sleep pattern. This prevalence of academic stress was also found in other study from medical colleges in Kolkata, India. ^17^. In their study, most of the students reported stress due to academic related factors which was greater among students who were not conversant with the local language. Similar significant association between higher levels of stress and sleep disturbance in college students have been shown in these following studies in which stress or anxiety affected quality of sleep of students.^18,19^ The reason for this can be accounted to the academic burden and constant pressure that the medical students face which is more than any other academic course in the country. However, in our current study, language was not a significant stress factor.

Further the academic performance of first assessment was significantly associated with change in sleeping pattern as compared to the other three assessments. Similar inference on significant association between sleep disturbances like insomnia and self-reported sleep quality and CGPA (Cumulative Grade Point Average) of medical and paramedical students were studied by Alquddah M et al. (2019)^20^. Another study^21^ suggested that students with poorer sleep quality are at a higher risk of impaired academic performance. The findings in our study can be attributed to the slacking attitude of the students in further assessments especially evident in students with lesser score than fifty percent score.

Similar significant association between academic score and duration of sleep in was evident in our three out of four assessments. There could be a direct association between academic score and duration of sleep or the less duration of sleep may make students more prone to stress or anxiety and affect academic performance indirectly. Hours of sleep acquired before exam time has been identified as a predictor of exam scores among medical students.^22^

As data on prevalence of SSB use among medical students were not available, we did not do apriori sample size calculation. However, taking 50% prevalence (as in pilot studies), 20% relative precision and α = 0.05, the minimum sample size required was 100.

### Strength and limitations of the study

The study had included good number of responses from the students of initial semesters of medical education and had adopted paperless data collection forms which helped in obtaining responses amidst lockdown due to Covid-19 pandemic. The sensitization of medical students to various coping strategies of stress will surely help them in future. The cross-sectional study design, lack of stratification of participants semester wise and recall bias had added to the limitation of this study. As this study was conducted from a single premier tertiary care teaching hospital, the generalizability of results could not be accomplished.

## Conclusion

Sleep has a very important impact on a person’s health. Medical students are no exception. Majority of students in our study had a change in sleeping pattern. Also, association between stress factors and change in sleeping pattern were observed and that the academic stress factors proved to be more significantly associated than social stress factors. The academic performance of students was also found to be associated with change in sleeping pattern and duration of sleep.

### Recommendations

There is an impending need to conduct various workshops focusing on stress management and awareness of importance of good quality sleep among medical students. There is a need to address these issues and provide the students with guidance and help to cope with them as students are the pillars of future health care.

## Supporting information

Supplementary Questionnaire file 1

STROBE checklist

## Data Availability

Data is available in the Supplementary Data File excel.

---

**Supplemental table 1.**
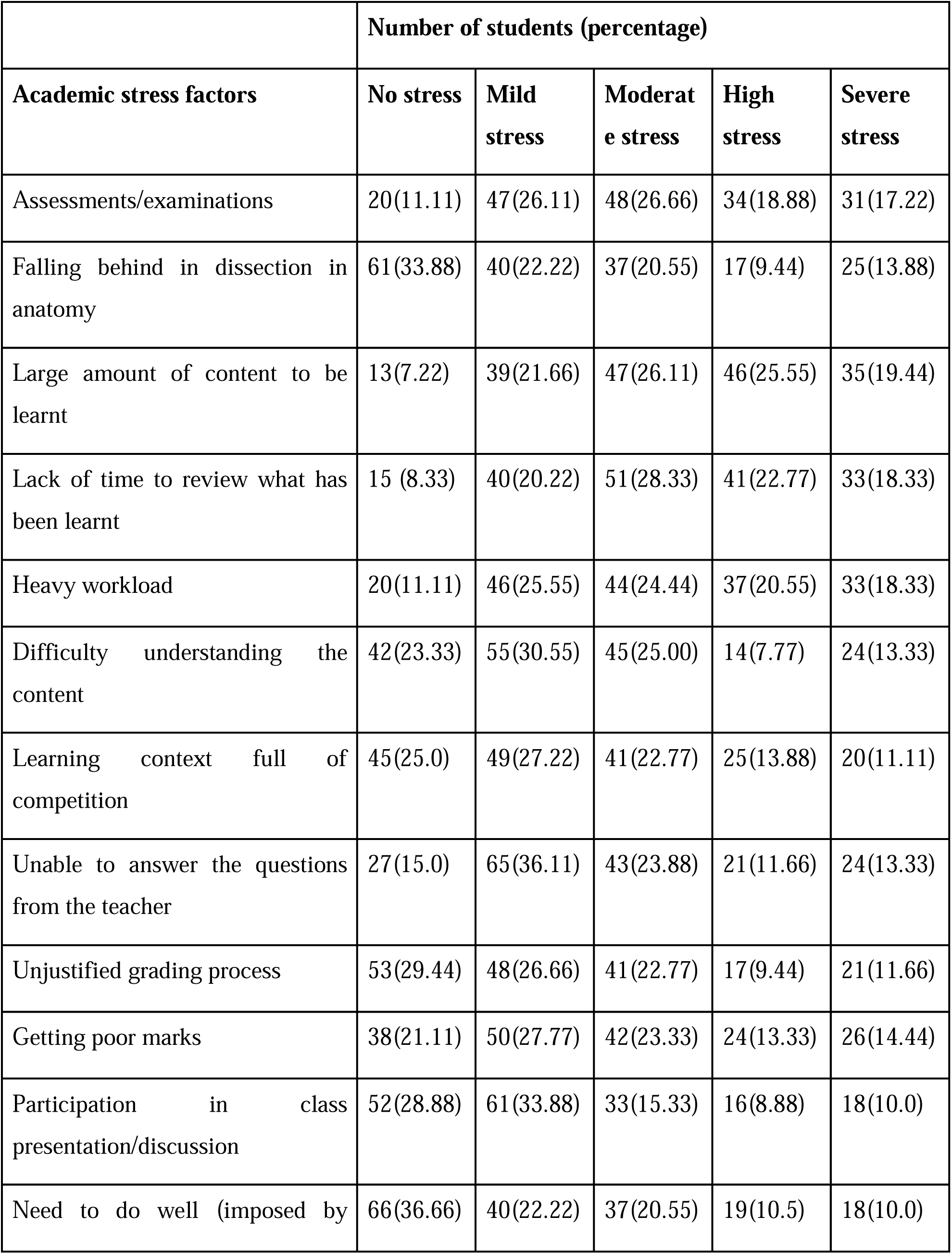
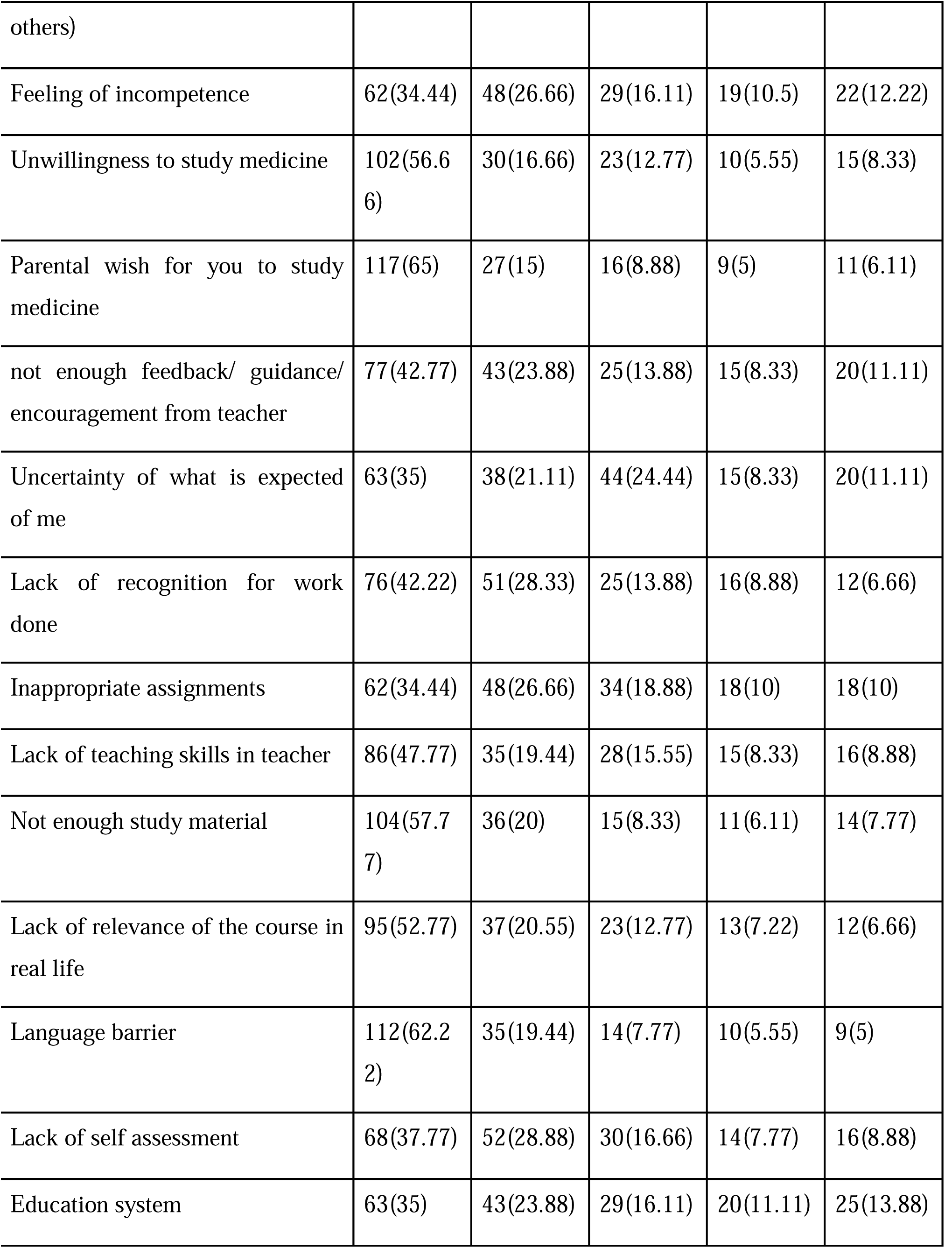
Response on academic stress factors.

**Supplemental table 2.** Response on social stress factors.

|  | Number of students (percentage) |  |  |  |  |
| --- | --- | --- | --- | --- | --- |
| **Social stress factors** | **No stress** | **Mild stress** | **Moderate stress** | **High stress** | **Severe stress** |
| Unable to answer questions from patients | 103(57.22) | 41(22.77) | 22(12.22) | 8(4.44) | 6(3.33) |
| Talking to patients about personal problems | 105(58.33) | 41(22.77) | 19(10.5) | 11(6.11) | 4(2.2) |
| Facing illness or death of patients | 97(53.88) | 41(22.77) | 27(15) | 8(4.44) | 7(3.88) |
| Frequent interruption of my work by others | 86(47.77) | 47(26.11) | 22(12.22) | 13(7.22) | 12(6.66) |
| Verbal or physical abuse by other students | 106(58.88) | 34(18.88) | 18(10) | 12(6.66) | 10(5.55) |
| Conflict with teachers and students | 99(55) | 29(16.11) | 31(17.22) | 10(5.55) | 11(6.11) |
| Lack of time for family | 68(37.77) | 54(30) | 27(15) | 18(10) | 13(7.22) |
| Lack of time for friends | 78(43.33) | 53(29.44) | 22(12.22) | 14(7.77) | 13(7.22) |
| Peer pressure | 92(51.11) | 34(18.88) | 37(20.55) | 9(5) | 8(4.44) |
| Problems with friends | 94(52.22) | 42(23.33) | 32(17.77) | 4(2.2) | 8(4.44) |
| Poor motivation to learn | 72(40) | 48(26.66) | 34(18.88) | 12(6.66) | 14(7.77) |
| Working with computer | 98(54.44) | 42(23.33) | 23(12.77) | 7(3.88) | 10(5.55) |
| Financial issues | 104(57.77) | 34(19.44) | 24(13.33) | 7(3.88) | 10(5.55) |
| Health issues | 91(50.55) | 42(23.33) | 30(16.66) | 6(3.33) | 11(6.11) |
| Challenge of living alone | 92(51.11) | 40(22.22) | 28(15.55) | 10(5.55) | 10(5.55) |
| Transportation issues | 103(57.22) | 36(20) | 23(12.77) | 8(4.44) | 10(5.55) |

**Supplemental table 3.**
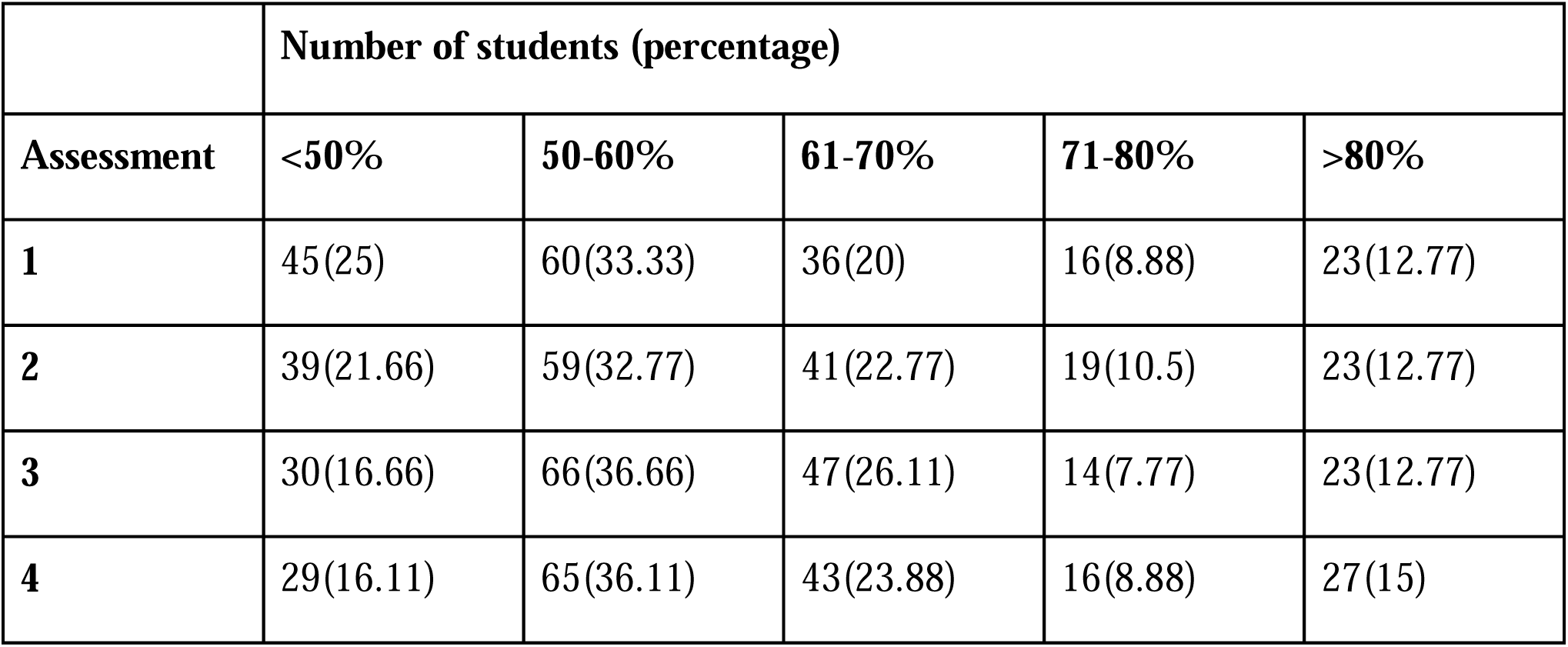
Academic record of participants.

## Additional files

1. Figure 1: Copying strategies
2. Supplementary Questionnaire File 1
3. Supplementary Data file 2
4. STROBE Checklist

## References

1. Corlateanu A, Covantev S, Botnaru V, et al. To sleep, or not to sleep – that is the question, for polysomnography. Breathe 2017; 13: 137–140.https://breathe.ersjournals.com/content/breathe/13/2/137.full.pdf

2. (Carskadon MA, Dement WC. Normal human sleep: an overview. In: Kryger MH, Roth T, Dement WC, editors. Principles and practice of sleep medicine. 4th ed. Philadelphia, PA: Elsevier Saunders; 2005. pp. 13–23. [Google Scholar]

3) Wolfson AR, Carskadon MA. Understanding adolescents sleep patterns and school performance:a critical appraisal. Sleep Med Rev 2003; 7: 491–506 [Google Scholar]

4) WHO technical meeting on sleep and health, Bonn Germany 22-24 January, 2004

5) Pagel, J F, Forister, N and Kwiatkowki, C. Adolescent sleep disturbance and. School performance : the confounding variable of socio economics. J Clin Sleep Med. 2007; 15:19–23. [Google Scholar]

6) Azad MC, Fraser K, Rumana N, Abdullah AF, Shahana N, Hanly PJ, Turin TC disturbances among medical students: a global perspective. J Clin Sleep Med. 2015;11(1):69–74. [Google Scholar]

7) AlFakhri L, Sarraj J, Kherallah S, Kuhail K, Obeidat A, Abu-Zaid A. Perceptions of pre-clerkship medical students and academic advisors about sleep deprivation and its relationship to academic performance: a crosssectional perspective from Saudi Arabia. BMC research notes. Dec 2015;8(1):740–748. [Google Scholar]

8) Dahlin M, Joneborg N, Runeson B. Stress and depression among medical students: a cross-sectional study. Med Educ. 2005;39:594–604. [PubMed] [Google Scholar]

9) Wilkinsos TJ, Gill DJ, Fitzjohn J, Palmer CL, Mulder RT. The impact on students of adverse experiences during medical school. Med Teach. 2006;28:129–35. [PubMed] [Google Scholar]

10) Ross S, Cleland J, Macleod MJ. Stress, debt and undergraduate medical performance. Med Educ. 2006;40:584–9. [PubMed] [Google Scholar]

11) Roberts J. Junior doctors’ years: training not education. BMJ. 1991;302:225–8. [PMC free article][PubMed] [Google Scholar]

12) Firth-Cozen J. Emotional distress in junior hospital doctors. BMJ. 1987;295:533–6. [PMC free article][PubMed] [Google Scholar]

13) Tyssen R, Vaglum P, Gronvold NT, Ekeberg O. The relative importance of individual and organizational factors for the prevention of job stress during internship: a nationwide and prospective study. Med Teach. 2005;27:27–31. [PubMed] [Google Scholar]

14) Willcock S, Daly MG, Tennant CC, Allard BJ. Burnout and psychiatric morbidity in new medical graduates. Med J Aust. 2004;181:357–60. [PubMed] [Google Scholar]

15) Hamed, H., Miskey, A., Alkurd, R., Ghazal, Z., Sami, R., Abduljaleel, N., & Shahwan, M. (2015). The effect of sleeping pattern on the academic performance of undergraduate medical students at Ajman University of Science and Technology. Journal of Pharmacy and Pharmaceutical Sciences, 4(4). H

16) Alsaggaf, Mohammed A., et al. “Sleep quantity, quality, and insomnia symptoms of medical students during clinical years: relationship with stress and academic performance.” Saudi medical journal 37.2 (2016): 173.

17) Gupta S, Choudhury S, Das M, Mondol A, Pradhan R. Factors causing stress among students of a medical college in Kolkata, India. Educ Health 2015;28:92–5

18) Lee SY, Wuertz C, Rogers R, Chen YP. Stress and sleep disturbances in female college students. Am J Health Behav. 2013;37:851–858. [PubMed] [Google Scholar]

19) Feng GS, Chen JW, Yang XZ. [Study on the status and quality of sleep-related influencing factors in medical college students] Zhonghua Liu Xing Bing Xue Za Zhi. 2005;26:328–331. Chinese. [PubMed] [Google Scholar]

20) Alqudah M, Balousha SAM, Al-Shboul O, Al-Dwairi A, Alfaqih MA, Alzoubi KH. Insomnia among Medical and Paramedical Students in Jordan: Impact on Academic Performance. Biomed Res Int. 2019 Oct 31;2019:7136906. doi: 10.1155/2019/7136906. PMID: 31781637; PMCID: PMC6875015.

21) Yeung WF, Chung KF, Cy Chan Tsleep-wake habits, excessive daytime sleepiness and academic performance among medical students in Hong Kong, biol Rhythm Res;2008;39:369–77.

22) Sitticharoon C, Srisuma S, Kanavitoon S, Summachiwakij S. Exploratory study of factors related to educational scores of first preclinical year medical students. Adv Physiol Educ. 2014;38:25–33. [PubMed] [Google Scholar]

