## Supplementary Questionnaire file 1 for "Evaluation of sleep pattern due to stress in undergraduate medical students and its impact on health and academic performance: A cross-sectional study from tertiary health center of Central India"

Name-

Time of sleeping

- Before 12 am
- 12 am to 2 am
- After 2 am

Duration of sleep

- <6 hours
- 6-8 hours
- >8 hours

Any change in sleeping pattern

- Present
- Absent

Any use of medication

- Present
- Absent

| **Academic stress factors** | | | | **Options** | | | | |
| --- | --- | --- | --- | --- | --- | --- | --- | --- |
|  | | | | No stress | Mild stress | Moderate stress | High stress | Severe stress |
| Tests/examinations | | | |  |  |  |  |  |
| Failing behind in dissection in anatomy | | | |  |  |  |  |  |
| Large amount of content to be learnt | | | |  |  |  |  |  |
| Lack of time to review what has been learnt | | | |  |  |  |  |  |
| Heavy workload | | | |  |  |  |  |  |
| Difficulty understanding the content | | | |  |  |  |  |  |
| Learning context full of competition | | | |  |  |  |  |  |
| Unable to answer the questions from the teacher | | | |  |  |  |  |  |
| Unjustified grading process | | | |  |  |  |  |  |
| Getting poor marks | | | |  |  |  |  |  |
| Participation in class presentation/discussion | | | |  |  |  |  |  |
| Need to do well imposed by others | | | |  |  |  |  |  |
| Feeling of incompetence | | | |  |  |  |  |  |
| Unwillingness to study medicine | | | |  |  |  |  |  |
| Parental wish for you to study | | | |  |  |  |  |  |
| Not enough feedback/ guidance and encouragement from teacher | | | |  |  |  |  |  |
| Uncertainty of what is expected of me | | | |  |  |  |  |  |
| Lack of recognition for work done | | | |  |  |  |  |  |
| Inappropriate assignments | | | |  |  |  |  |  |
| Lack of teaching skills in teacher | | | |  |  |  |  |  |
| Not enough study material | | | |  |  |  |  |  |
| Lack of relevance of the course in real life | | | |  |  |  |  |  |
| Language barrier | | | |  |  |  |  |  |
| Lack of self assessment | | | |  |  |  |  |  |
| Education system | | | |  |  |  |  |  |

| **Social stress factors** | | | | **Options** | | | | |
| --- | --- | --- | --- | --- | --- | --- | --- | --- |
|  | | | | No stress | Mild stress | Moderate stress | High stress | Severe stress |
| Unable to answer questions from patients | | | |  |  |  |  |  |
| Talking to patients about personal problems | | | |  |  |  |  |  |
| Facing illness or death of patients | | | |  |  |  |  |  |
| Frequent interruption of my work by others | | | |  |  |  |  |  |
| Verbal or physical abuse by other students | | | |  |  |  |  |  |
| Conflict with teachers and students | | | |  |  |  |  |  |
| Lack of time for family | | | |  |  |  |  |  |
| Lack of time for friends | | | |  |  |  |  |  |
| Peer Pressure | | | |  |  |  |  |  |
| Problems with friends | | | |  |  |  |  |  |
| Poor motivation to learn | | | |  |  |  |  |  |
| Working with computer | | | |  |  |  |  |  |
| Financial issues | | | |  |  |  |  |  |
| Health issues | | | |  |  |  |  |  |
| Challenge of living alone | | | |  |  |  |  |  |
| Transportation issues | | | |  |  |  |  |  |

|  | **Academic record of participants (in last tests**) | | | | |
| --- | --- | --- | --- | --- | --- |
|  | Below 50 | 50-60 | 61-70 | 71-80 | Above 80 |
| Test 1 |  |  |  |  |  |
| Test 2 |  |  |  |  |  |
| Test 3 |  |  |  |  |  |
| Test 4 |  |  |  |  |  |

| **Coping strategies to control stress** | | | | **Options** | | | | |
| --- | --- | --- | --- | --- | --- | --- | --- | --- |
|  | | | | Never done | I have not been doing this | U have done this a little bit | I have been doing this a medium amount | I have been doing this a lot |
| Time management helps me | | | |  |  |  |  |  |
| Good Balanced diet reduce stress | | | |  |  |  |  |  |
| Deep breathing exercise and yoga helps me | | | |  |  |  |  |  |
| Watching TV/ comedy shows | | | |  |  |  |  |  |
| I involve in religious coping reframing | | | |  |  |  |  |  |
| I plan things ahead | | | |  |  |  |  |  |
| Study helps me to relax | | | |  |  |  |  |  |
| Meditation and muscle relaxation helps me | | | |  |  |  |  |  |
| Exercise / sports helps me | | | |  |  |  |  |  |
| Music/ book reading (hobby) helps me to relax | | | |  |  |  |  |  |
| Good sleep relaxes me | | | |  |  |  |  |  |
| Talking to family members/ friends helps me | | | |  |  |  |  |  |
| I need emotional support | | | |  |  |  |  |  |
| I predict and accept the stress as a challenge | | | |  |  |  |  |  |
| Self blame myself for stress | | | |  |  |  |  |  |
| I go in denial | | | |  |  |  |  |  |
| I show my anger and anxiety | | | |  |  |  |  |  |
